# Socioeconomic and Regional Disparities in Postnatal Care Utilization in Bangladesh: Evidence from the Bangladesh Demographic and Health Survey 2022

**DOI:** 10.1101/2025.10.07.25337546

**Authors:** Sumaiya Shafayet Supti, Salma Sultana, Md. Muddasir Hossain Akib, Priom Saha, Margia Yesmin, Bikash Pal

## Abstract

Maternal and neonatal mortality in Bangladesh remains high, particularly in rural areas where access to skilled postnatal care (PNC) is limited. This study assessed urban-rural disparities in PNC utilization using data from the 2022 Bangladesh Demographic and Health Survey. A sample of 4,844 ever-married women aged 15–49 with a recent live birth (3,249 rural; 1,595 urban) was analyzed. Outcomes included received any PNC, timely PNC within two days, and PNC from trained providers. Socio-demographic, reproductive, and healthcare determinants are analyzed using survey adjusted weighted logistic regression. Overall, 76.6% of women received PNC, 70.4% within 48 hours, and 24.7% from skilled providers. Utilization was consistently higher in urban than rural areas. Education, wealth, antenatal visits, and maternal employment were positively associated with PNC use, with women attending four or more antenatal visits showing nearly twice the odds of receiving timely and skilled PNC. Conversely, higher birth order and regional disparities, particularly in Rangpur, were linked to reduced skilled care. Chattogram showed comparatively higher coverage. Persistent inequities in PNC utilization, with rural women lagging behind in both timeliness and access to skilled providers indicates targeted interventions addressing education, socio-economic inequality, and service availability are critical to improving maternal and newborn health outcomes in Bangladesh.

## Introduction

Maternal and neonatal mortality (MNM) remain pressing global health concerns, despite progress in maternal and child health over recent decades. Maternal deaths are categorized broadly as direct or indirect, with most resulting from complications such as severe postpartum hemorrhage, infections, prolonged labor, and unsafe abortions [1,2]. Neonatal deaths are caused frequently by sepsis, perinatal asphyxia, and prematurity or low birth weight [3]. Importantly, a large proportion of these deaths occur during the immediate postnatal period, particularly within the first 48 hours after delivery, highlighting the importance of skilled care and timely postnatal interventions [4–7]. These outcomes are preventable through high-quality maternal and newborn care, making MNM reduction a global health priority.

Globally, the scale of the problem is stark. Every 11 seconds, a woman or newborn dies, most from preventable causes [8]. In 2023, an estimated 260,000 maternal deaths occurred worldwide about 712 deaths daily [9]. While maternal mortality declined by 40% between 2000 and 2023, about 92% of these deaths still occur in low- and middle-income countries (LMICs), where maternal mortality ratios (MMR) are on average 15 times higher than in high-income settings [1,10–12]. The Sustainable Development Goal (SDG) framework underscores this issue through indicators such as 3.1.1 (MMR) and 3.1.2 (proportion of births attended by skilled health personnel) [12]. Despite global gains in skilled birth attendance, coverage remains uneven, with only 53% of deliveries attended by skilled health personnel in 2017 [6].

The situation is especially critical in LMICs, where resource constraints, weak health systems, and inequitable access to care amplify risks. Inadequate antenatal services, limited skilled attendance at delivery, and poor utilization of postnatal care (PNC) remain key drivers of maternal and neonatal deaths [13–15]. Rural populations are disproportionately affected due to poverty, lower literacy, cultural practices, and limited healthcare infrastructure [16–20]. In such contexts, timely and appropriate care depends heavily on family- and community-level practices, with the absence of skilled personnel at delivery or during the first day of life significantly increasing neonatal mortality risk [17,21].

Bangladesh has achieved notable progress in reducing maternal mortality, with its MMR declining from 523 in 2000 to 115 per 100,000 live births in 2023 representing a 79% reduction [22]. However, disparities persist. About 46% of births still occur at home, rising to 50% in rural areas [23], where only a minority are attended by trained professionals [24,25]. Rural women also report higher barriers to healthcare access compared to urban women [26], and both maternal and child mortality remain higher in rural areas [27]. Although the proportion of births attended by skilled professionals rose to about 70% by 2022 [28,29], projections suggest Bangladesh’s MMR in 2030 may still range between 150 and 200 per 100,000 live births [30], exceeding national targets [29]. These challenges highlight persistent inequities in maternal and newborn health service coverage.

Despite the improved availability of maternal health services, significant gaps remain in postnatal care utilization, particularly in the timely delivery of PNC within 48 hours and PNC by trained providers. Moreover, limited information exists on specific maternal and newborn care practices in rural Bangladesh, where health outcomes are most precarious. Understanding the barriers and enabling factors at the community level is essential for designing effective, culturally appropriate interventions [3,17,18].

This study addresses these gaps by examining urban-rural disparities in three critical maternal health outcomes-receipt of any PNC, timely PNC within two days after delivery, and PNC provided by trained health professionals. By analyzing individual- and community-level determinants, the study aims to identify service coverage gaps, guide policymakers and program planners, and inform strategies to increase awareness, promote early care-seeking, and expand access to skilled maternal health services. The findings will contribute to national and global goals of reducing maternal and neonatal morbidity and mortality, particularly in underserved rural communities of Bangladesh.

## Materials and methods

This study utilized data from the most recent Bangladesh Demographic and Health Survey (BDHS) 2022, a nationally representative cross-sectional survey that collects comprehensive information on demographic trends and key health indicators among Bangladeshi mothers. Ever married women aged 15–49 who had given birth in the past three years were interview for collecting data on the most recent birth. After excluding incomplete cases, the weighted sample for this study comprised 4,844 women, including 3,249 from rural areas and 1,595 from urban areas (Fig 2). Additional information on survey protocols and datasets can be accessed via the DHS website: https://dhsprogram.com/data/available-datasets.cfm.

**Fig 1:**
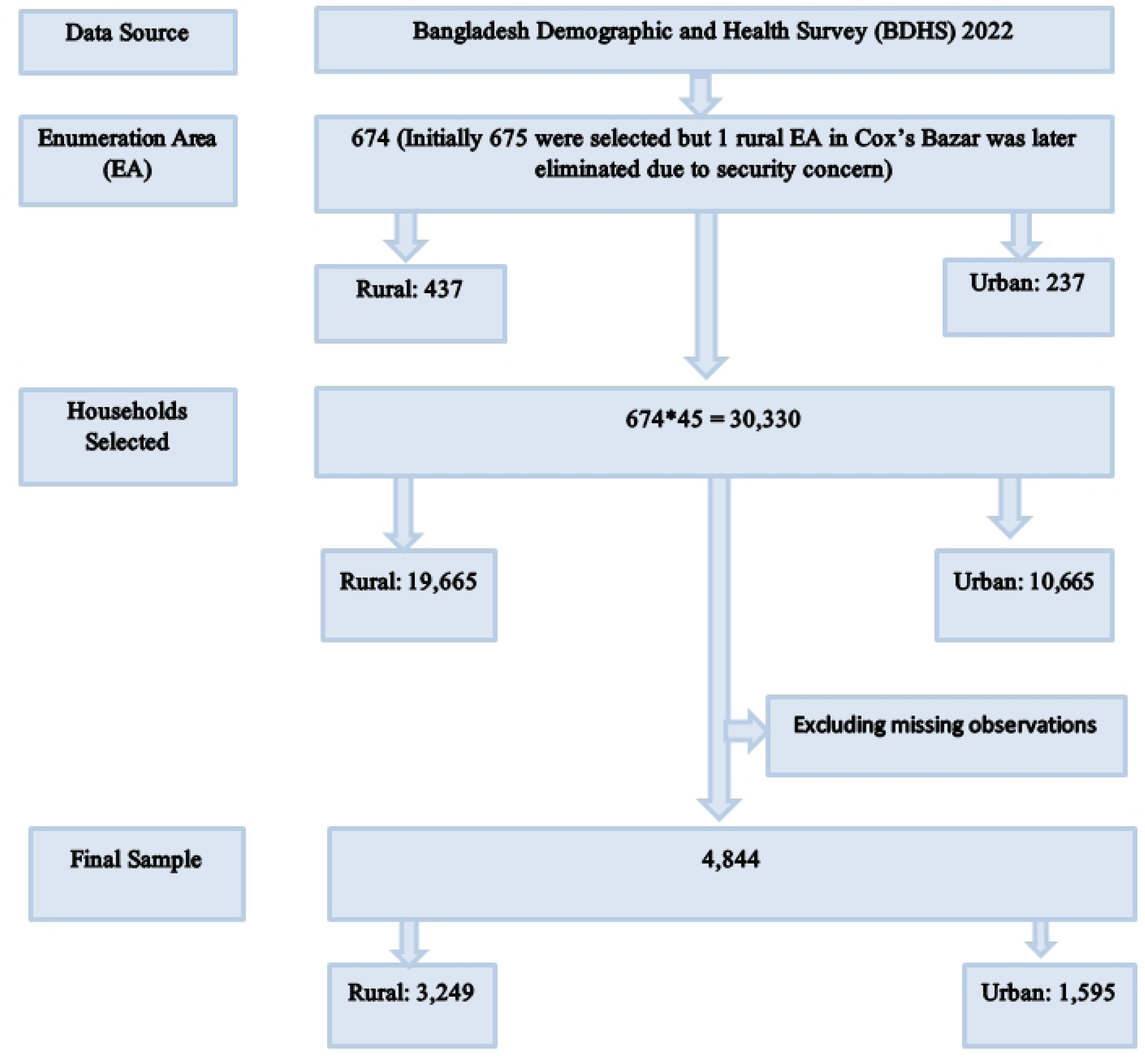
Flowchart of sample selection form BDHS 2022 dataset.

**Fig 2:**
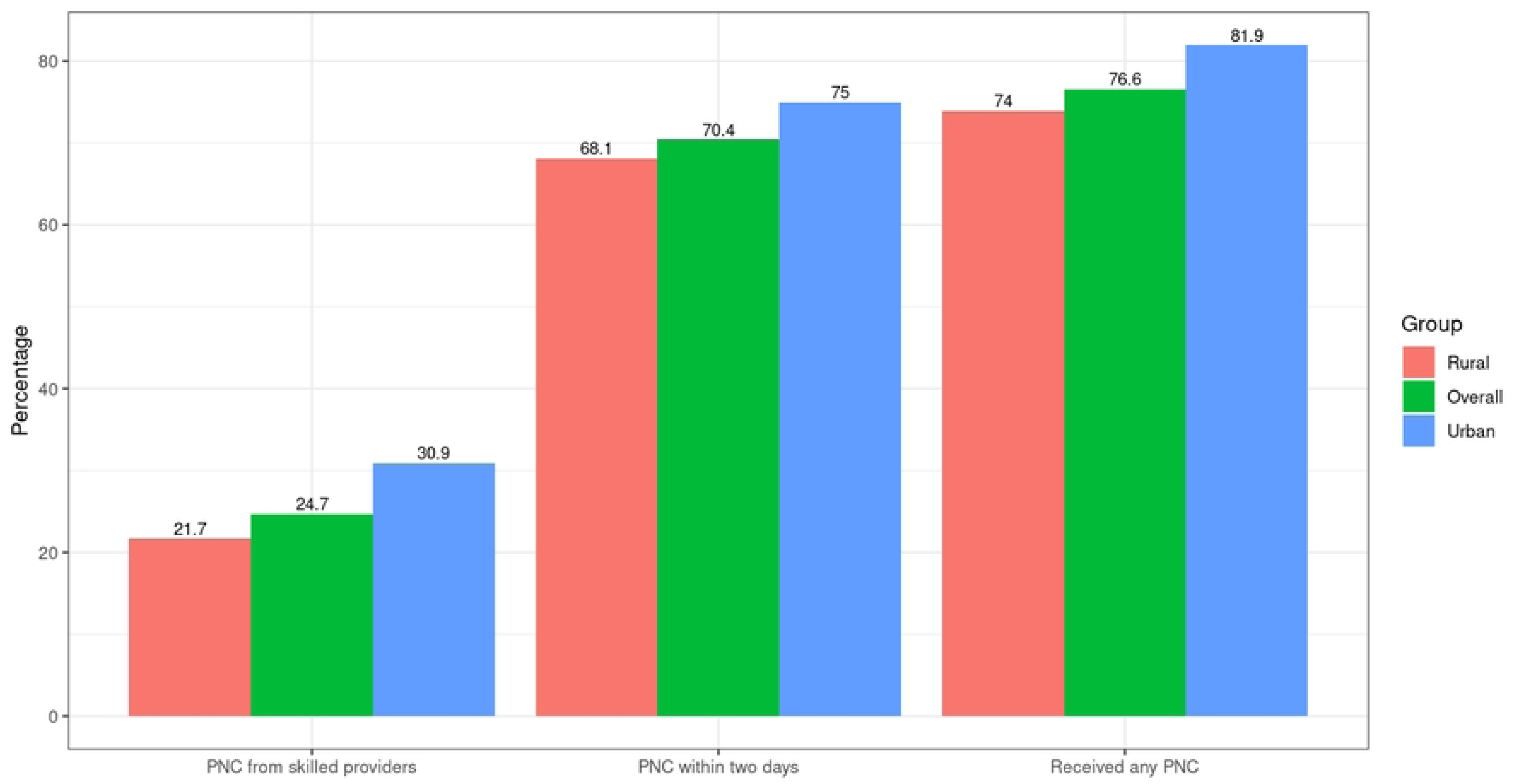
Utilization of Postnatal Care (PNC) Services by Residence in Bangladesh

## Outcome variables

This study examined three primary outcome variables related to PNC among women aged 15–49 years who had a live birth in the three years preceding the survey [31]. Utilization of postnatal care services captured whether the respondent received any form of postnatal care within 24 hours to 42 days following delivery, with responses recorded as “yes” for women who utilized postnatal care within this period and “no” for those who did not. Given that the highest risk of maternal and neonatal mortality occurs shortly after childbirth, the timing of the first PNC visit was categorized into two groups, “yes” for women who utilized postnatal care within the first two days after delivery and “no” for those who utilized postnatal care after two days. Health-seeking behavior for postnatal services was categorized based on the type of provider, where care received from a trained provider such as a qualified doctor, nurse, midwife, paramedic, family welfare visitor, community skilled birth attendant, community health care provider, health assistant, family welfare assistant, trained traditional birth attendant, or NGO worker was coded as “trained health care provider,” and services obtained from untrained traditional birth attendants, unqualified doctors, or others were categorized as “untrained health care provider” [32]. Responses were recorded as “yes” if the respondent received care from a trained health care provider and “no” if the care was provided by an untrained health care provider.

## Explanatory variables

This study considered a range of explanatory variables encompassing socio-demographic, reproductive, and healthcare-related characteristics to explore factors associated with PNC utilization [31,33]. Respondent and partner characteristics included the mother’s age, categorized “as less than 20 years”, “20-34 years”, and “35-49 years” [34,35], and age at first birth, grouped into “less than 16 years”, “16-21 years”, and “22 or more years”. Educational attainment for both respondents and their husbands was divided into two categories, “no/primary” and “secondary or higher.” The respondent’s current working status was dichotomized as “yes” or “no,” while the husband’s occupation was categorized into “job,” “business,” and “others.” Household characteristics included the wealth index, grouped into quintiles (poorest, poorer, middle, richer, richest) [31,33], and media exposure, which was classified as “yes” or “no” based on access to at least one form of mass media (television, radio, or newspaper) [36,37]. The number of household members was also recorded (less than 4, 4-6, and 7 and above), and type of toilet facility was categorized as “flash,” “pit,” and “other” [33]. Reproductive and child-related variables included birth order (first, second or third, fourth or more), sex of the most recent child (male, female), religion (Islam, others) [38] and the age of the child at the time of the survey (less than 7 months, 7–24 months, more than 24 months). Whether the mother experienced the death of any child was also included as a binary variable as “yes” or “no”. Healthcare-related factors included antenatal care (ANC) attendance, categorized as “less than 4” and “4 and above” visits. Whether the mother had experienced fever recently was captured as a binary variable as “yes” or “no”. Perceived barriers to accessing healthcare were coded as “at least one problem” or “no problem,” based on common difficulties such as cost, distance, or lack of permission. Delivery-related characteristics covered the place of delivery grouped into “respondent’s home,” “public hospital,” “private facility,” and “NGO and others” and mode of delivery, categorized into “normal vaginal delivery (NVD)” and “caesarean section.” Geographical factors included the respondent’s place of residence (urban, rural) and administrative division (Barisal, Chattogram, Dhaka, Khulna, Mymensingh, Rajshahi, Rangpur, Sylhet).

## Statistical methods

Firstly, descriptive statistics, including frequency distributions and percentages, were employed to summarize the characteristics of the study population. Bivariate analyses were then conducted to explore associations between postnatal care (PNC) utilization, considering timing and trained provider, and selected socio-demographic and healthcare-related variables. The Pearson chi-square test was applied to identify variables significantly associated with PNC utilization, which were subsequently considered for multivariable analysis. Variables showing a significant association with the three outcome variables at the 5% level were selected for inclusion in the multivariable analysis [38]. Finally, to account for the cluster sampling design of the BDHS 2022, where women are nested within clusters, a survey-weighted, cluster-adjusted multivariable logistic regression model was employed to examine the relationship between parental education and other covariates with the utilization of skilled postnatal care services [39]. The **svyset** command in STATA was applied during both the bivariate and multivariable analyses [39,40]. The results of the regression analyses were presented as adjusted odds ratios (AORs) with 95% confidence intervals (CIs), and statistical significance was set at a p-value of less than 0.05. All analyses were conducted using R and STATA 14.0.

## Results

The study examined urban-rural differences in postnatal care (PNC) utilization in Bangladesh and identified key determinants across three outcomes: receipt of any PNC, PNC within two days of delivery, and PNC from skilled providers.

Fig 2 illustrates the distribution of the Postnatal services which shows noteworthy disparities highlighting higher rates in urban areas and consistently lower rates in rural areas. As presented in **Table 1**, overall, PNC coverage was higher among urban mothers compared with rural mothers. In the descriptive analysis, about 75% of urban mothers reported receiving any PNC compared with 60% in rural areas. Timely PNC (within two days) was reported by 63% of urban and 50% of rural mothers, while skilled PNC was received by 58% of urban versus 44% of rural mothers. Utilization was also higher among mothers with secondary or higher education (urban: 80%; rural: 65%) compared to those with no or primary education (urban: 62%; rural: 52%). Similar gradients were observed by household wealth: the richest urban mothers had the highest coverage of any PNC (over 85%), while only 45% of the poorest rural mothers received care. Mothers who attended four or more antenatal care (ANC) visits had substantially greater PNC coverage (urban: 82%; rural: 68%) compared with those with fewer visits (urban: 55%; rural: 41%). Regional variations were evident, with Khulna and Chattogram reporting higher levels of PNC, while Rangpur and Sylhet lagged behind.

**Table 1.**
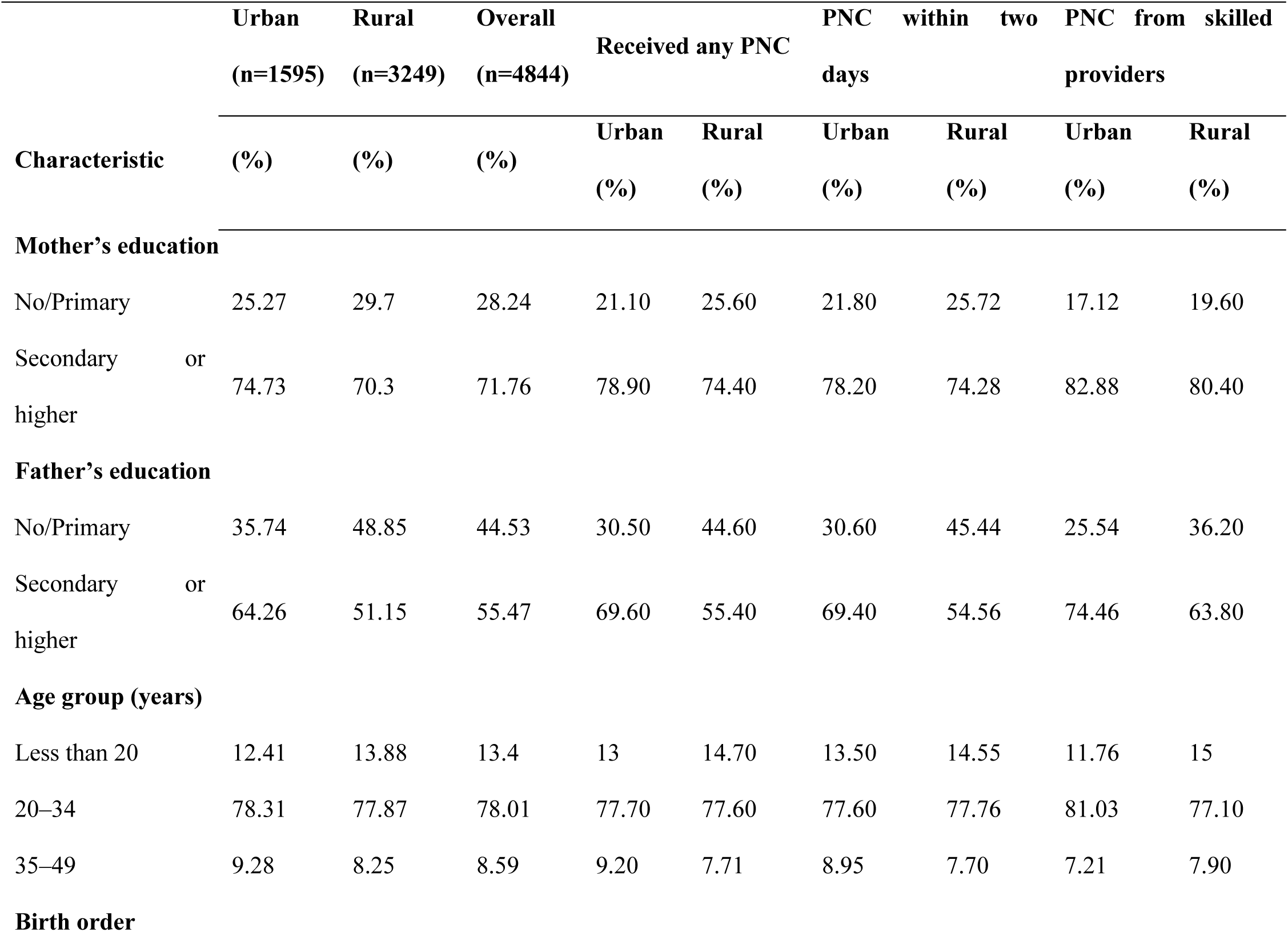
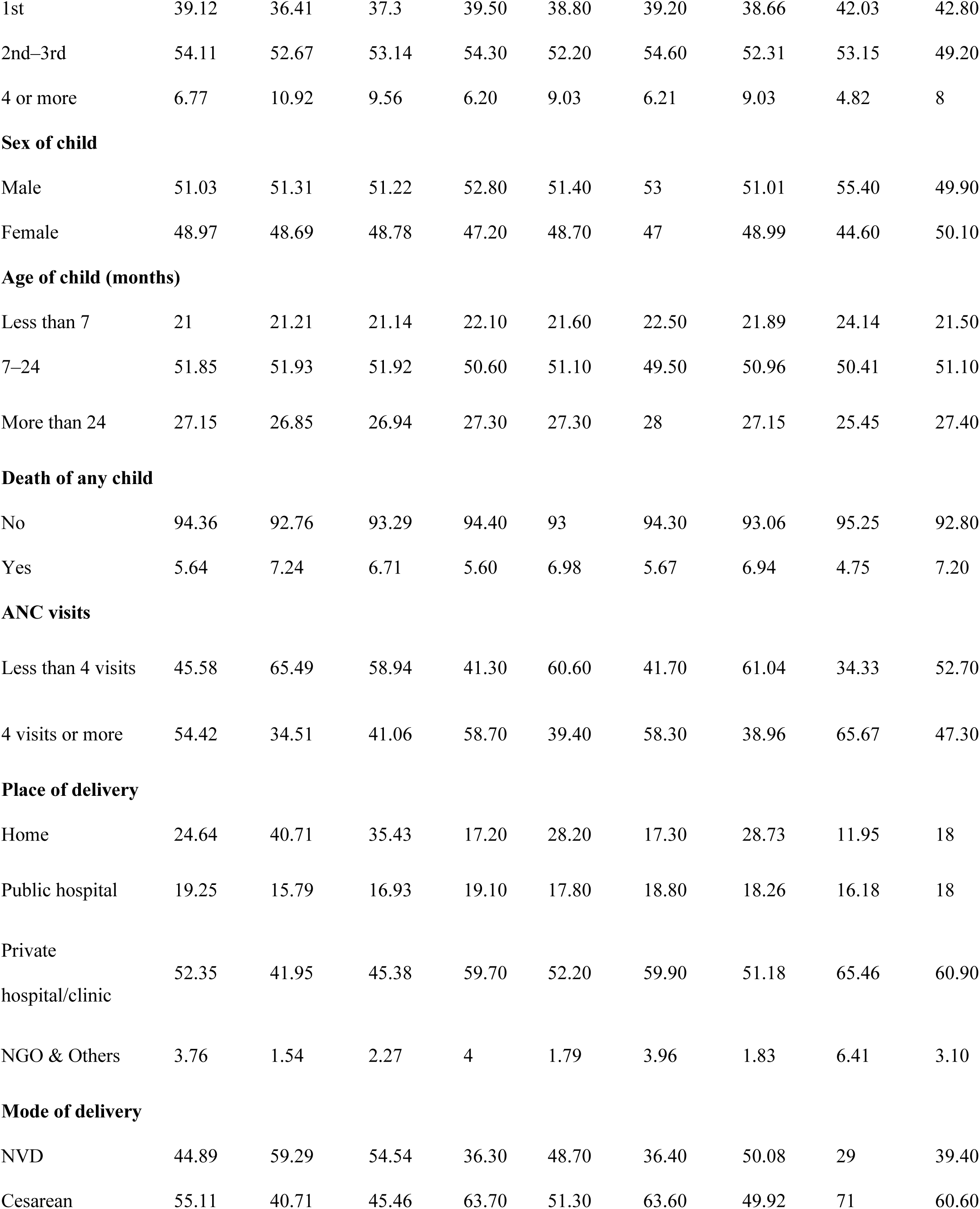
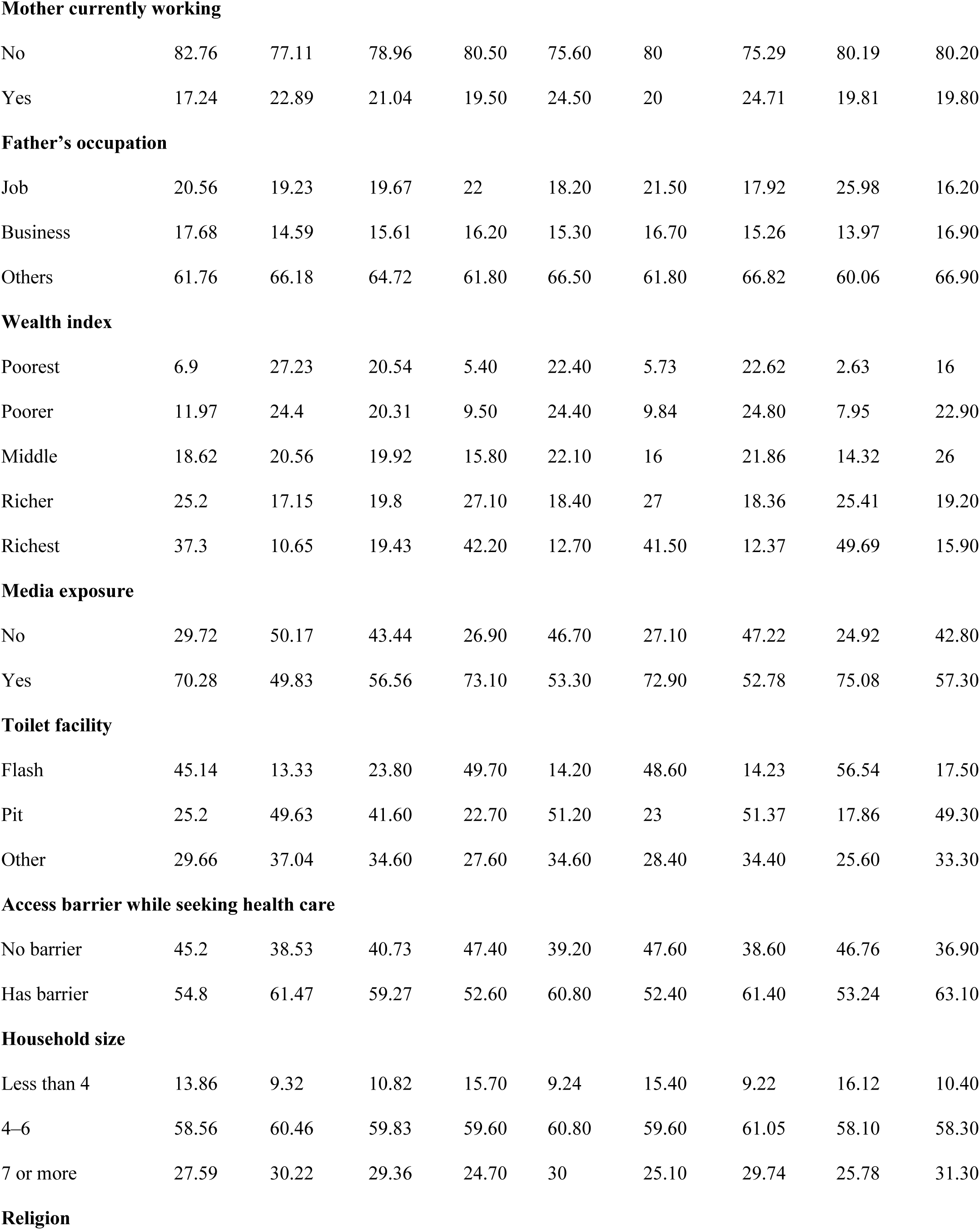
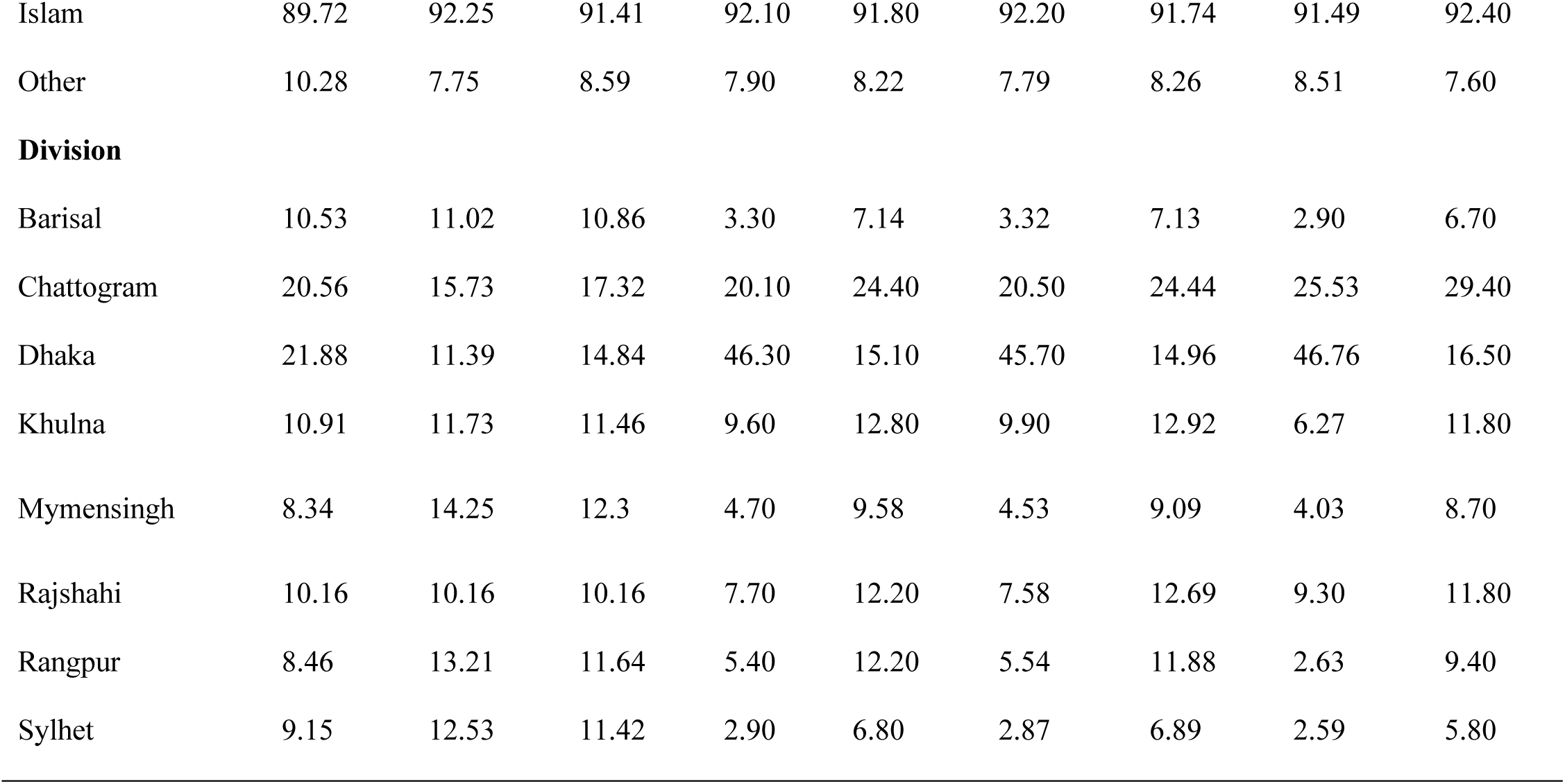
Univariate and Bivariate distribution of postnatal care utilization among women by socio-demographic and maternal characteristics (%).

As shown in **Table 2**, the multivariable analyses provided further support for these patterns. Education emerged as a strong determinant: urban mothers with secondary or higher education had higher odds of receiving any PNC (AOR = 1.63) and skilled PNC (AOR = 1.36), while in rural areas the effect was smaller but significant (AOR = 1.22 and 1.33, respectively). Father’s education was more influential in rural settings, where higher paternal education increased the odds of any PNC (AOR = 1.32, p < 0.01) and skilled PNC (AOR = 1.41, p < 0.01). Birth order showed a negative association in rural areas. Compared with first births, mothers with four or more children were significantly less likely to receive any PNC (AOR = 0.64, p < 0.01) or timely PNC (AOR = 0.68, p < 0.01). No such gradient was seen among urban mothers. ANC visits remained one of the strongest predictors across all settings. Mothers with four or more ANC visits were about twice as likely to receive any PNC in both urban (AOR = 2.01) and rural (AOR = 2.11) areas, with similarly strong effects for timely and skilled PNC. Maternal employment influenced rural outcomes: working mothers were more likely to receive any PNC (AOR = 1.63, p < 0.01) and timely PNC (AOR = 1.54, p < 0.001), while no significant effect was observed in urban areas. Wealth-related disparities were marked. Rural mothers in the richest quintile had significantly higher odds of receiving any PNC (AOR = 1.86, p < 0.01), while those in the middle and richest quintiles also had higher odds of skilled PNC (AOR = 1.57 and 1.40, respectively). In urban areas, the richest mothers were much more likely to receive skilled PNC (AOR = 2.44). Other household factors, including media exposure, toilet facility, and access barriers, showed no consistent associations. Religion influenced rural outcomes, with non-Muslim mothers more likely to receive any PNC (AOR = 1.38, p < 0.05) and timely PNC (AOR = 1.35, p < 0.05). Regional differences were prominent. Compared to Barisal, mothers in Khulna had higher odds of any PNC in both urban (AOR = 1.65) and rural (AOR = 1.62) areas. Urban mothers in Chattogram were more likely to receive skilled PNC (AOR = 2.21, p < 0.01), while those in Rangpur were less likely (AOR = 0.46, p < 0.05). Rural mothers in Rangpur also had lower odds of receiving any PNC (AOR = 0.66) and skilled PNC (AOR = 0.71).

**Table 2.**
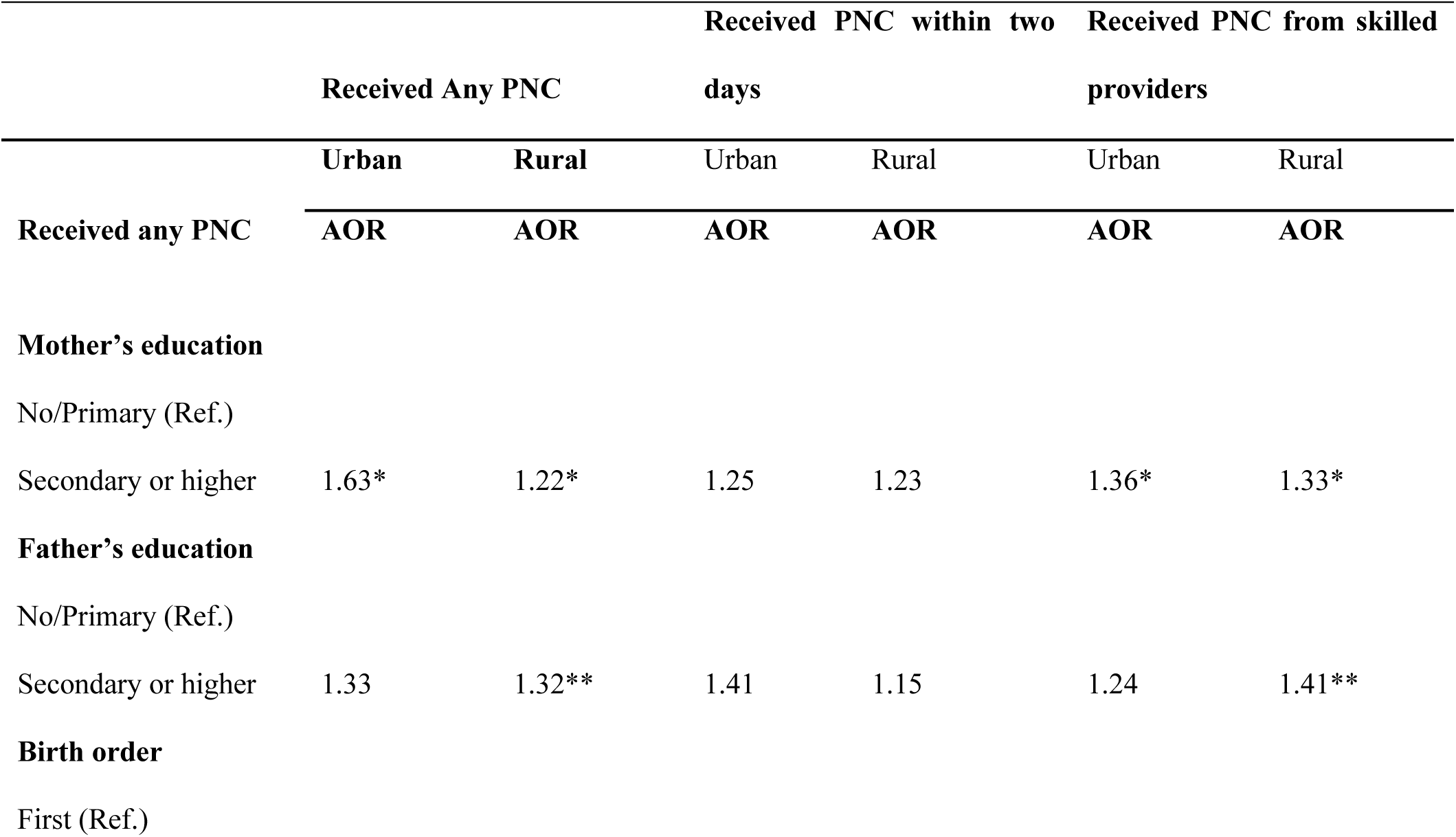
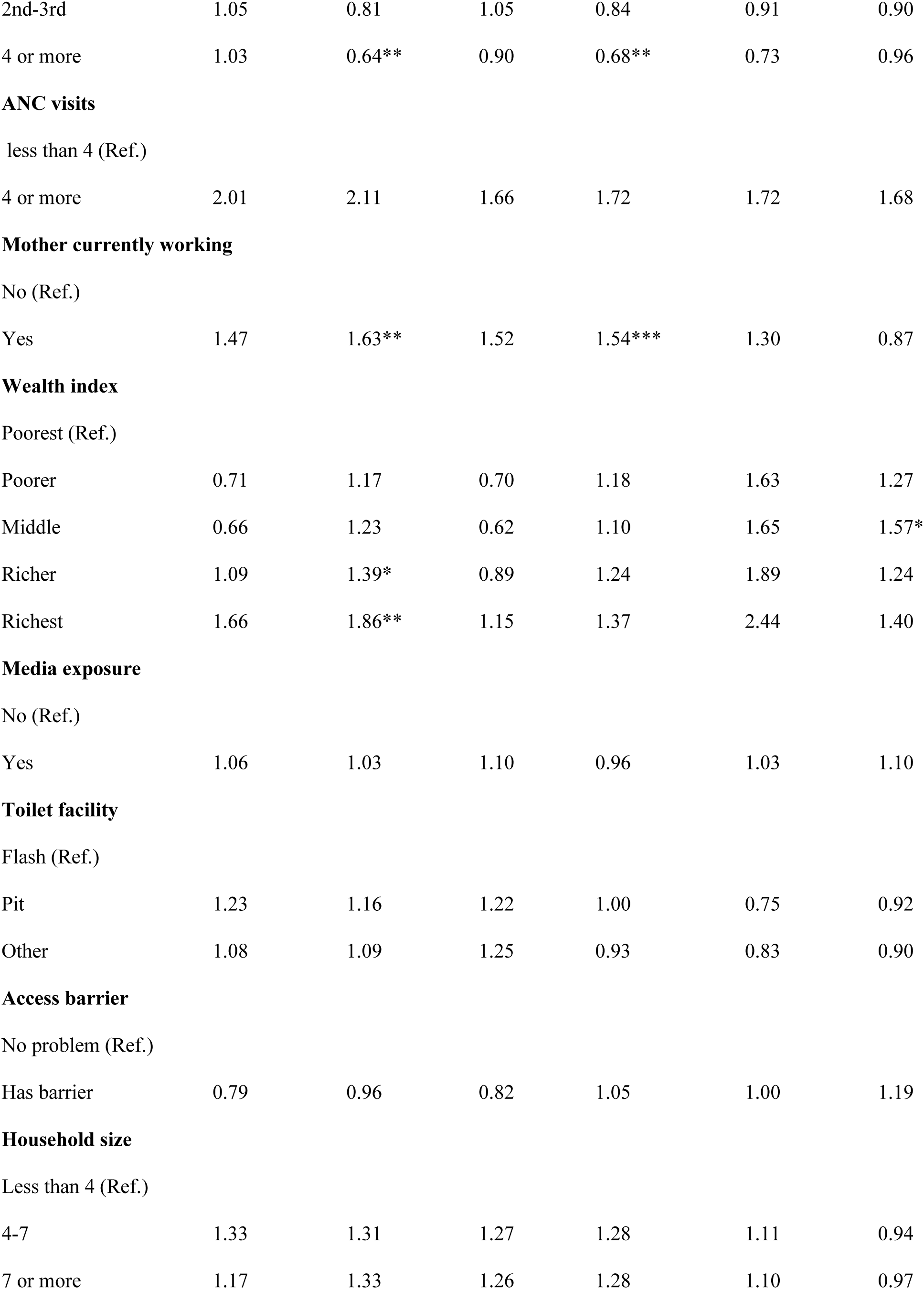
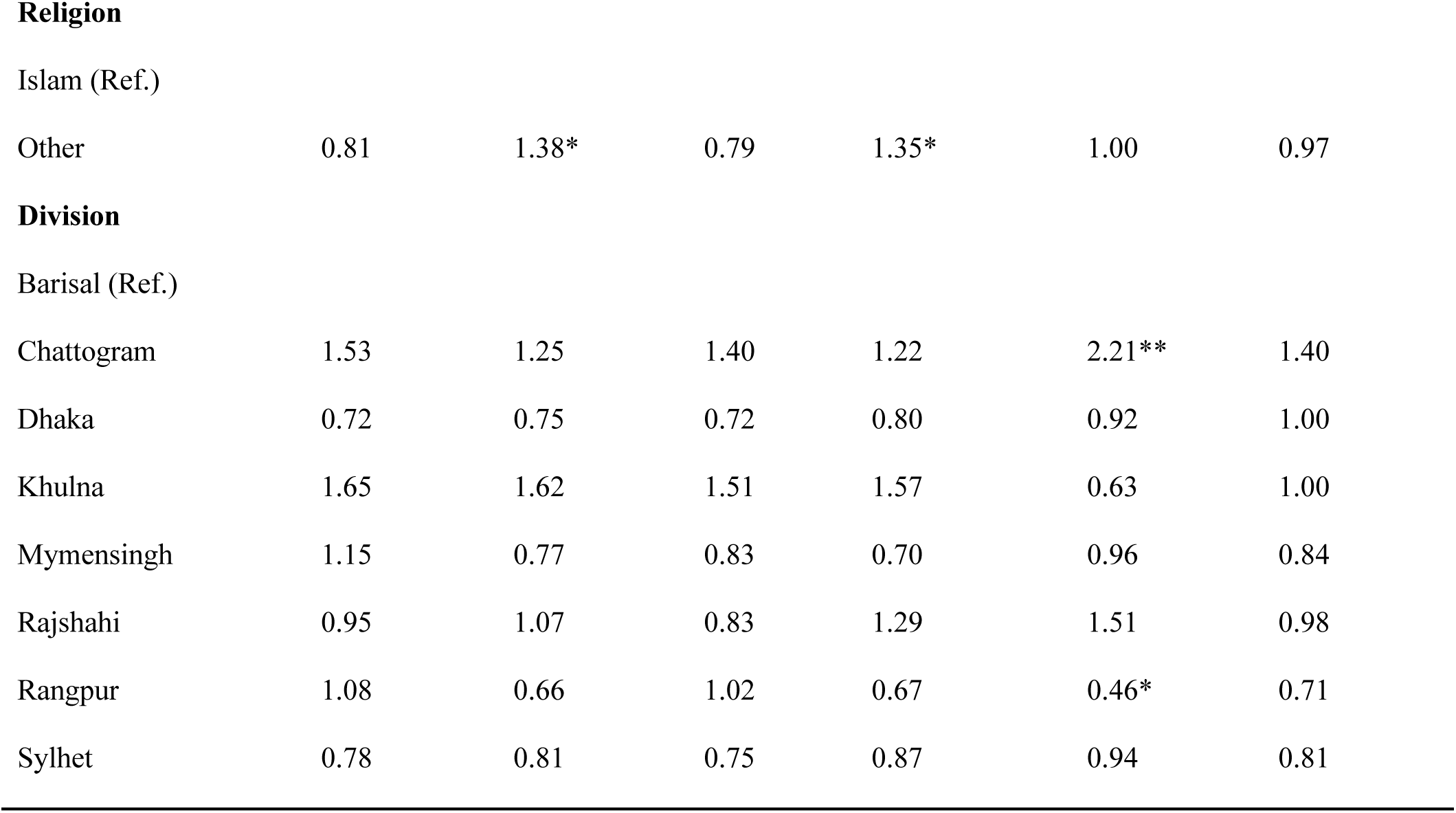
The factors associated with receiving any PNC, receiving PNC within two working days and receiving PNC from skilled providers based on multivariable analysis.

|  | Received Any PNC |  | Received PNC within two days |  | Received PNC from skilled providers |  |
| --- | --- | --- | --- | --- | --- | --- |
|  | Urban | Rural | Urban | Rural | Urban | Rural |
| Received any PNC | AOR | AOR | AOR | AOR | AOR | AOR |
| **Mother's education** |  |  |  |  |  |  |
| No/Primary (Ref.) |  |  |  |  |  |  |
| Secondary or higher | 1.63* | 1.22* | 1.25 | 1.23 | 1.36* | 1.33* |
| **Father's education** |  |  |  |  |  |  |
| No/Primary (Ref.) |  |  |  |  |  |  |
| Secondary or higher | 1.33 | 1.32** | 1.41 | 1.15 | 1.24 | 1.41** |
| **Birth order** |  |  |  |  |  |  |
| First (Ref.) |  |  |  |  |  |  |
| 2nd-3rd | 1.05 | 0.81 | 1.05 | 0.84 | 0.91 | 0.90 |
| 4 or more | 1.03 | 0.64** | 0.90 | 0.68** | 0.73 | 0.96 |
| **ANC visits** |  |  |  |  |  |  |
| less than 4 (Ref.) |  |  |  |  |  |  |
| 4 or more | 2.01 | 2.11 | 1.66 | 1.72 | 1.72 | 1.68 |
| **Mother currently working** |  |  |  |  |  |  |
| No (Ref.) |  |  |  |  |  |  |
| Yes | 1.47 | 1.63** | 1.52 | 1.54*** | 1.30 | 0.87 |
| **Wealth index** |  |  |  |  |  |  |
| Poorest (Ref.) |  |  |  |  |  |  |
| Poorer | 0.71 | 1.17 | 0.70 | 1.18 | 1.63 | 1.27 |
| Middle | 0.66 | 1.23 | 0.62 | 1.10 | 1.65 | 1.57* |
| Richer | 1.09 | 1.39* | 0.89 | 1.24 | 1.89 | 1.24 |
| Richest | 1.66 | 1.86** | 1.15 | 1.37 | 2.44 | 1.40 |
| **Media exposure** |  |  |  |  |  |  |
| No (Ref.) |  |  |  |  |  |  |
| Yes | 1.06 | 1.03 | 1.10 | 0.96 | 1.03 | 1.10 |
| **Toilet facility** |  |  |  |  |  |  |
| Flash (Ref.) |  |  |  |  |  |  |
| Pit | 1.23 | 1.16 | 1.22 | 1.00 | 0.75 | 0.92 |
| Other | 1.08 | 1.09 | 1.25 | 0.93 | 0.83 | 0.90 |
| **Access barrier** |  |  |  |  |  |  |
| No problem (Ref.) |  |  |  |  |  |  |
| Has barrier | 0.79 | 0.96 | 0.82 | 1.05 | 1.00 | 1.19 |
| **Household size** |  |  |  |  |  |  |
| Less than 4 (Ref.) |  |  |  |  |  |  |
| 4-7 | 1.33 | 1.31 | 1.27 | 1.28 | 1.11 | 0.94 |
| 7 or more | 1.17 | 1.33 | 1.26 | 1.28 | 1.10 | 0.97 |
| Other | 0.81 | 1.38* | 0.79 | 1.35* | 1.00 | 0.97 |
| Chattogram | 1.53 | 1.25 | 1.40 | 1.22 | 2.21** | 1.40 |
| Dhaka | 0.72 | 0.75 | 0.72 | 0.80 | 0.92 | 1.00 |
| Khulna | 1.65 | 1.62 | 1.51 | 1.57 | 0.63 | 1.00 |
| Mymensingh | 1.15 | 0.77 | 0.83 | 0.70 | 0.96 | 0.84 |
| Rajshahi | 0.95 | 1.07 | 0.83 | 1.29 | 1.51 | 0.98 |
| Rangpur | 1.08 | 0.66 | 1.02 | 0.67 | 0.46* | 0.71 |
| Sylhet | 0.78 | 0.81 | 0.75 | 0.87 | 0.94 | 0.81 |

## District-wise distribution of PNC utilization from skilled providers

The geospatial variations in PNC utilization from skilled providers across Bangladesh are reflected from the spatial mapping (Fig 3, Fig 4, Fig 5). Overall, PNC utilization exceeded 45% in districts such as Natore, Narshingdi, Feni, and Chattogram, while it fell below 15% in Nawabganj, Patuakhali, Dinajpur, Rangpur, Magura, and Munshiganj. Urban areas showed generally higher skilled PNC utilization, with several districts, including Natore, Thakurgaon, Rajbari, Bagura, Narshingdi, Habiganj, Khagrachhari, Rangamati, and Chattogram, reporting 50% or more. In contrast, Nilphamari, Kishoreganj, and Barguna showed 0% skilled PNC utilization, with some districts showing missing data due to the absence of urban clusters. In rural areas, utilization remained above 40% in Natore, Narshingdi, Feni, and Chattogram but dropped below 15% in Dinajpur, Nawabganj, Rajbari, Magura, Khagrachhari, Munshiganj, Gazipur, Sylhet, and Patuakhali.

**Fig 3:**
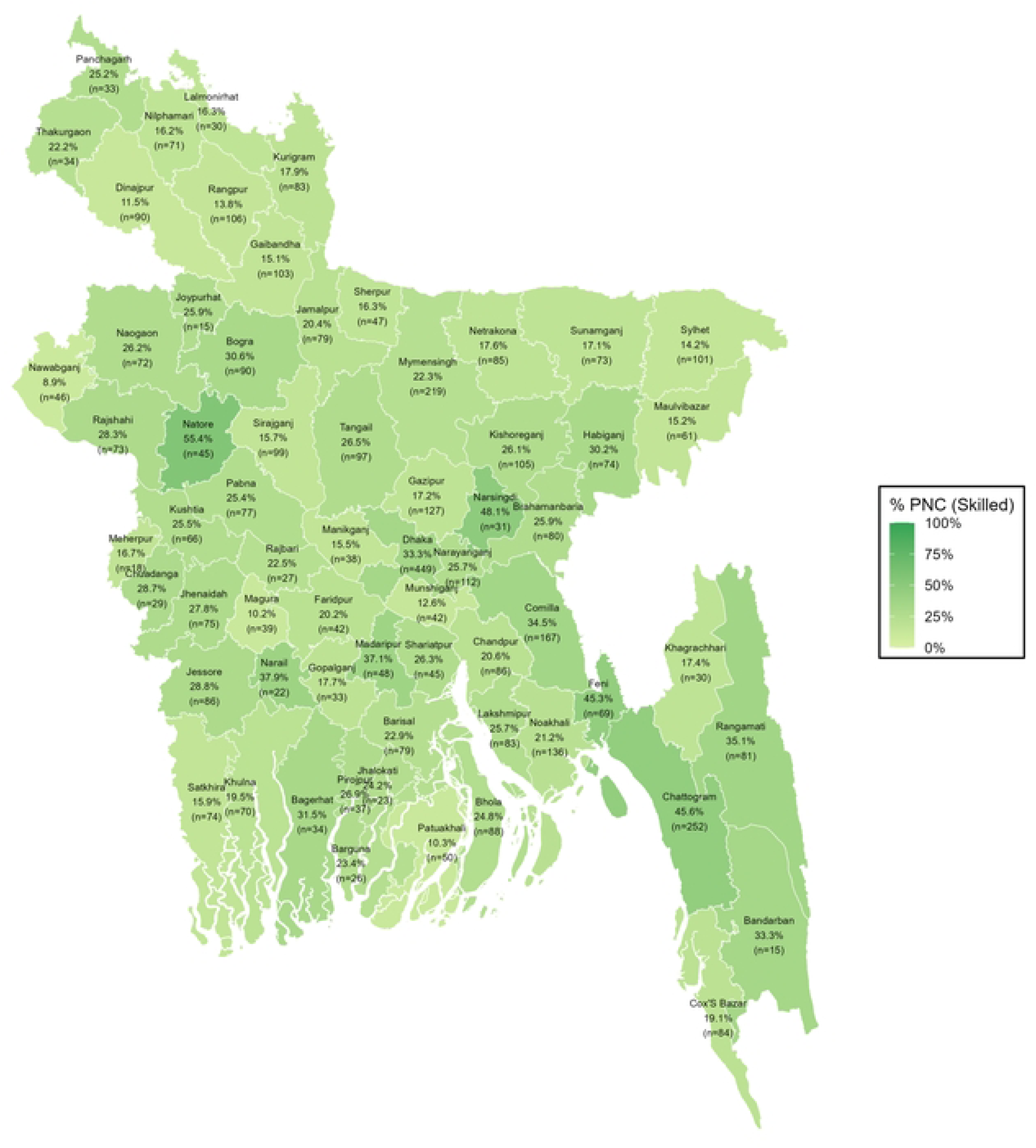
District-wise distribution of PNC utilization from skilled providers among all women

**Fig 4:**
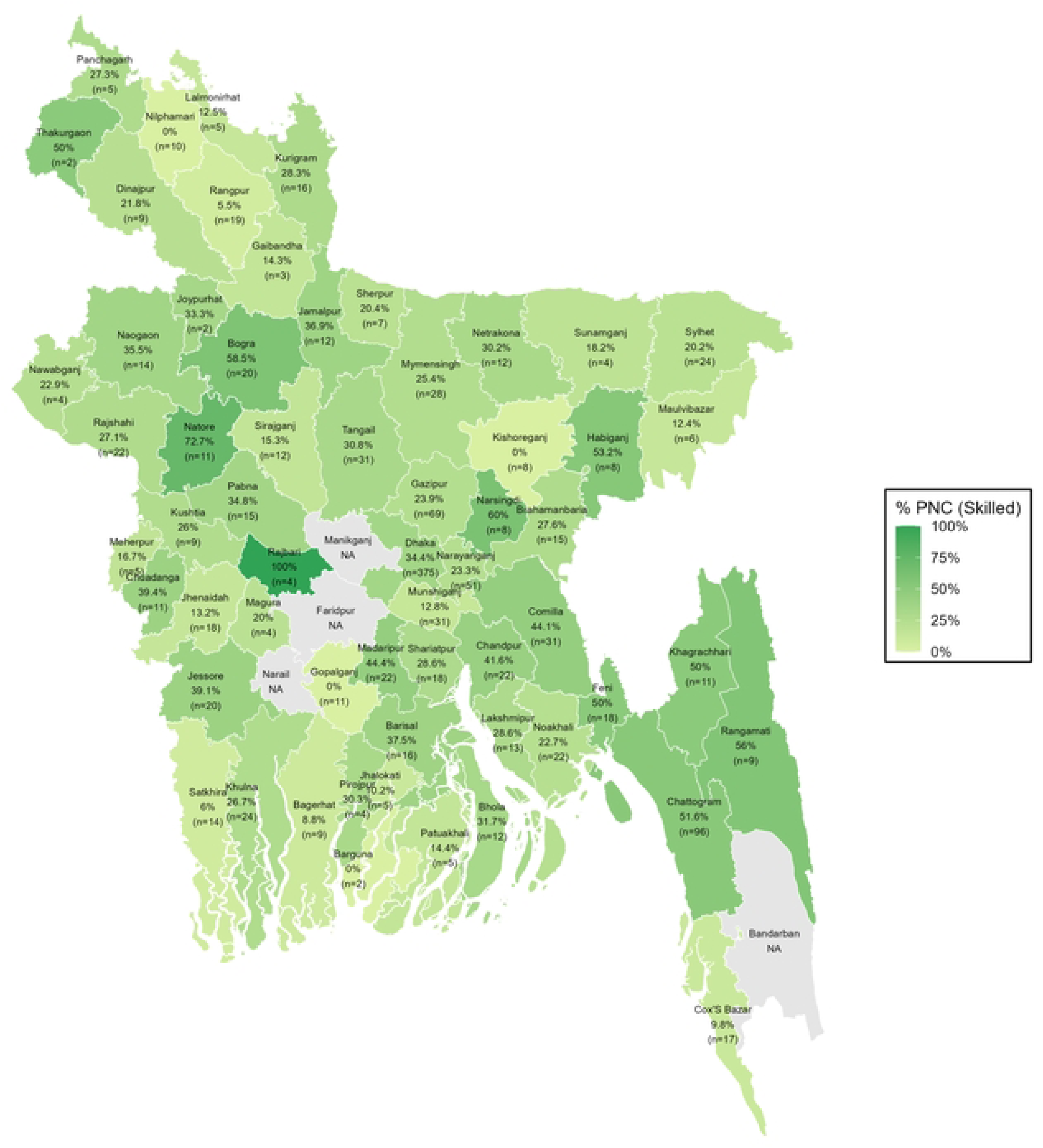
District-wise distribution of PNC utilization from skilled providers among urban women

**Fig 5:**
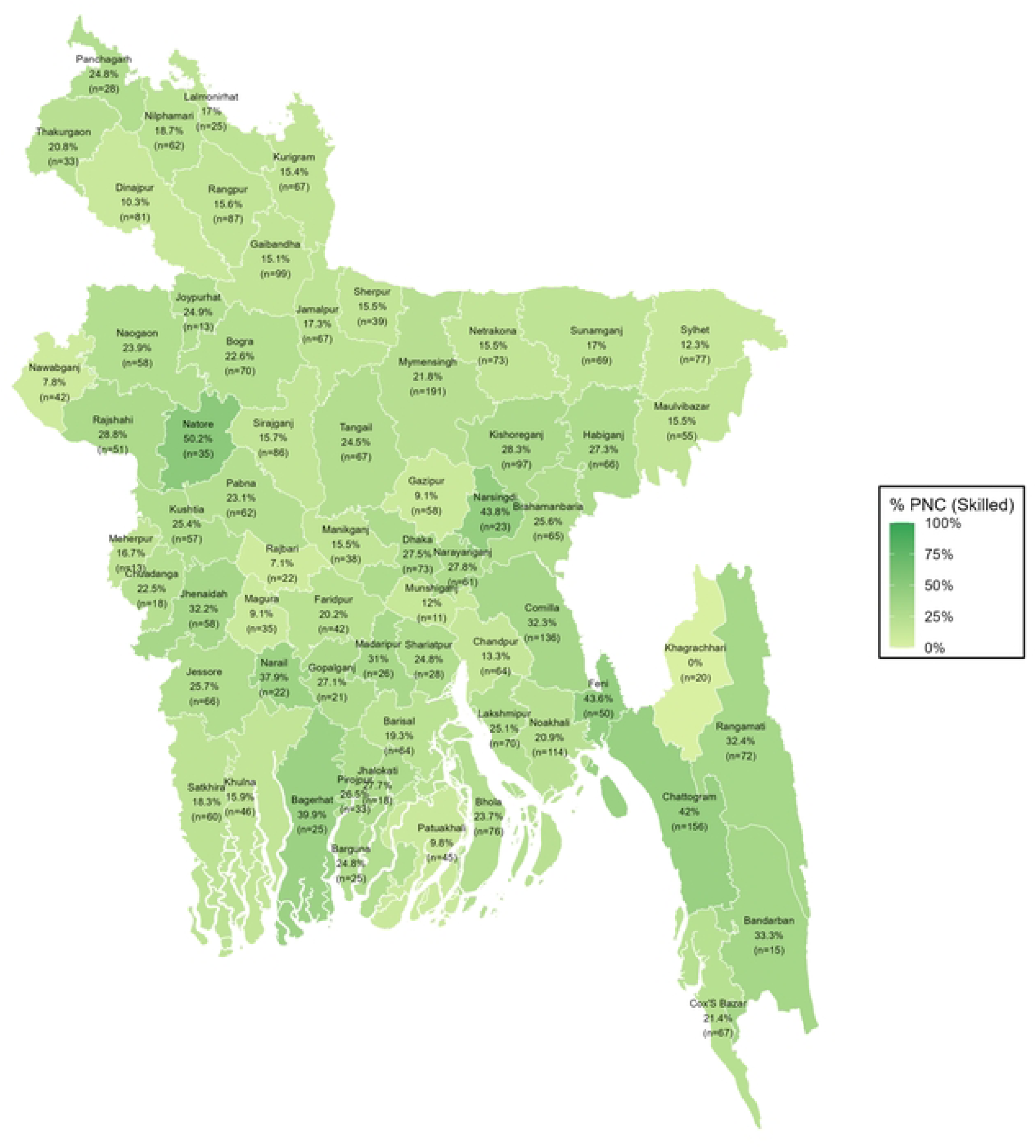
District-wise distribution of PNC utilization from skilled providers among rural women

In summary, both bivariate and multivariable results highlight education, ANC utilization, maternal employment, wealth, and region as the most influential factors shaping PNC uptake, with rural mothers consistently facing greater barriers to timely and skilled care. These findings demonstrate sustained inequities in postnatal service use across Bangladesh.

## Discussion

This study examined patterns and determinants of PNC utilization in Bangladesh, focusing on urban-rural disparities across three outcomes: receipt of any PNC, timely PNC within two days, and PNC from skilled providers. The findings revealed substantial inequalities, with rural women consistently lagging behind urban counterparts in utilization, timeliness, and access to skilled care. While overall coverage has improved over time-rising from 29% in 2011 [18] to 55% in 2022 [32] for timely PNC the proportion receiving skilled postnatal care remains low, particularly in rural areas, where only about 20% of women reported skilled attendance [4,5]. Compared with regional peers, Bangladesh still demonstrates modest PNC uptake; countries such as India (65%) [41], Myanmar (68%) [42], Nepal (59%) [43] and Indonesia (78.5%) [44] report higher coverage, highlighting the need for continued efforts to strengthen maternal health services.

Education emerged as one of the most influential factors shaping PNC utilization. Mothers with secondary or higher education and those with similarly educated partners had greater odds of receiving any PNC and skilled PNC, confirming the role of education in shaping health-seeking behaviors [31,33,40]. Educated women are more likely to access information, make autonomous decisions, and utilize health services, while paternal education particularly influenced rural women, reflecting male-dominated household decision-making in these contexts [45–48]. ANC attendance was the most consistent predictor, with women completing four or more visits being twice as likely to receive PNC, timely checks, and skilled care, supporting the continuum of maternal care observed in South Asia and sub-Saharan Africa [40,49–56].

In addition, socioeconomic status, maternal employment, and wealth were significant determinants. Working women and those from wealthier households were more likely to obtain timely PNC, particularly in rural areas, likely due to increased autonomy, financial capacity, and exposure to health information [57–60]. Wealth gradients were pronounced in rural settings, where economic status determined early ANC attendance, facility delivery, and skilled PNC use, while urban disparities were less marked [61–63]. Birth order demonstrated an inverse relationship with PNC utilization, with higher-order births reducing the likelihood of receiving care, particularly in rural areas [28,33,57,64,65]. This pattern may reflect resource constraints, reduced perceived risk, or reliance on maternal experience from previous births.

Urban–rural comparisons brought to light structural and contextual inequalities. Rural women faced barriers such as limited transportation, long travel distances, and reliance on untrained providers, contributing to lower utilization and timeliness of PNC [66–69]. Regional disparities further accentuated inequities: rural women in Khulna had relative advantages in timely PNC, while urban mothers in Chattogram accessed skilled providers more frequently, with Rangpur lagging in both urban and rural settings. Maternal employment had a stronger association in rural areas, enhancing general care-seeking, though it did not guarantee access to skilled providers. Religion also influenced PNC uptake, with Muslim women more likely to access services compared with women practicing traditional faiths [70].

These findings showcase the interrelation of education, wealth, ANC attendance, maternal employment, birth order, and regional factors in shaping PNC utilization. They highlight the need for targeted interventions that improve access to skilled postnatal services, particularly in rural areas, while addressing socio-economic and educational disparities. Strengthening community outreach, promoting awareness of early PNC, and ensuring availability of trained providers are essential for reducing maternal and neonatal morbidity and mortality. Urban-rural gaps persist, but evidence suggests that improving women’s education, economic empowerment, and health system capacity can substantially enhance postnatal care coverage and equity [57,71,72,72,73].

## Strengths and limitations

This study used nationally representative data from BDHS 2022 and applied precise statistical analysis, including survey design adjustments. However, the cross-sectional nature of the data limits causal inference, and recall bias may have affected reporting of PNC service use. Despite these limitations, the results provide important insights into current PNC utilization patterns and highlight areas requiring policy attention.

## Conclusion

This study reveals persistent urban–rural disparities in postnatal care utilization in Bangladesh. While overall PNC coverage has improved, rural women consistently lag behind urban counterparts in receiving any PNC, timely checks within two days, and care from skilled providers. Maternal and paternal education, attendance of four or more antenatal visits, maternal employment, and household wealth emerged as strong positive determinants of PNC utilization, whereas higher birth order and regional differences reduced access. Urban and rural contexts showed distinct patterns: paternal education and wealth had stronger effects in rural areas, while maternal education was more influential in urban settings. These findings underscore the need for targeted interventions, including educational initiatives, expansion of antenatal services, enhanced access to skilled providers, and policies that address socio-economic and geographic inequities. Strengthening these efforts could improve timely and skilled postnatal care, ultimately contributing to better maternal and newborn health outcomes across Bangladesh.

## Author Contributions

**Conceptualization:** Md. Muddasir Hossain Akib; **Data curation:** Sumaiya Shafayet Supti; **Formal analysis:** Sumaiya Shafayet Supti, Salma Sultana; **Methodology:** Sumaiya Shafayet Supti; **Writing – original draft:** Sumaiya Shafayet Supti, Salma Sultana; **Writing – review & editing:** Priom Saha, Md. Muddasir Hossain Akib.

## Data Availability

The dataset is publicly available at DHS website: https://dhsprogram.com/data/available-datasets.cfm.

## Acknowledgments

We sincerely acknowledge the Demographic and Health Surveys (DHS) Program for providing access to the dataset used in this study. We also extend our gratitude to the National Institute of Population Research and Training (NIPORT) for conducting the Bangladesh Demographic and Health Survey (BDHS) 2022, which made this research possible.

